# Detection of SARS-CoV-2 in pets living with COVID-19 owners diagnosed during the COVID-19 lockdown in Spain: A case of an asymptomatic cat with SARS-CoV-2 in Europe

**DOI:** 10.1101/2020.05.14.20101444

**Authors:** Ignacio Ruiz-Arrondo, Aránzazu Portillo, Ana M. Palomar, Sonia Santibáñez, Paula Santibáñez, Cristina Cervera, José A. Oteo

**Author notes:** Corresponding author. E-mail address (José A. Oteo).

## Abstract

During April-May 2020, the presence of respiratory syndrome coronavirus 2 (SARS-CoV-2) in pets living with coronavirus disease 2019 (COVID-19) owners was analyzed. From 23 pets, a cat without clinical symptoms showed positive results for SARS-CoV-2 in oropharyngeal swab using three RT-qPCR assays (negative rectal swab). SARS-CoV-2 was not detected in the remaining pets. Our finding suggests that cats may act as asymptomatic dispersers of SARS-CoV-2, although viral transmission from animals to humans seems unlikely.

## Background

The natural origin of severe acute respiratory syndrome coronavirus 2 SARS-CoV-2), responsible for the current coronavirus disease 2019 (COVID-19) pandemic, seems derived from bats (spillover). Pangolins, snakes, turtles, hamsters or yaks have been considered as potential intermediate hosts before spreading to humans, but this still remains unclear [1]. According to WHO (May 8, 2020), there is no evidence that dogs and/or cats can disseminate SARS-CoV-2 and act as source of human infection [2]. Nevertheless, in any new or emerging disease and even more if there are ‘knowledge gaps’ in the epidemiology, to assess the potential for domestic transmission through pets (especially dogs and cats) is relevant [3](Our aim was to evaluate, at an early stage of COVID-19 pandemic, the state of infection and in consequence the potential role of transmission of SARS-CoV-2 from companion animals (especially dogs and cats) under a one health approach in La Rioja (northern Spain).

## Methods

From April 8-May 4, 2020, a total of 23 asymptomatic mammalian pets under quarantine(8 cats, 1 guinea pig, 2 rabbits and 12 dogs) from 17 households with confirmed human cases of COVID-19 infection diagnosed at the Hospital Universitario San Pedro (Logroño, Spain) were included in our study (Table). All residences were located in La Rioja (North of Spain). Institutional review board approval for this study was obtained from the Ethical Committee of Research with medicines from La Rioja (CEImLAR), reference number P.I. 419. Two samples per animal (oropharyngeal and rectal swabs) were collected and preserved at 4°C in 2 mL Dulbecco’s Modified Eagle Medium (DMEM) with penicillin (100 units/mL) and streptomycin (100 μg/mL) (Sigma-Aldrich, Manheim, Germany) up to the arrival (<5 h) to the Special Pathogens Laboratory-Center of Rickettsiosis and Arthropod-Borne Diseases-Center of Biomedical Research from La Rioja (CRETAV-CIBIR).

**Table.**
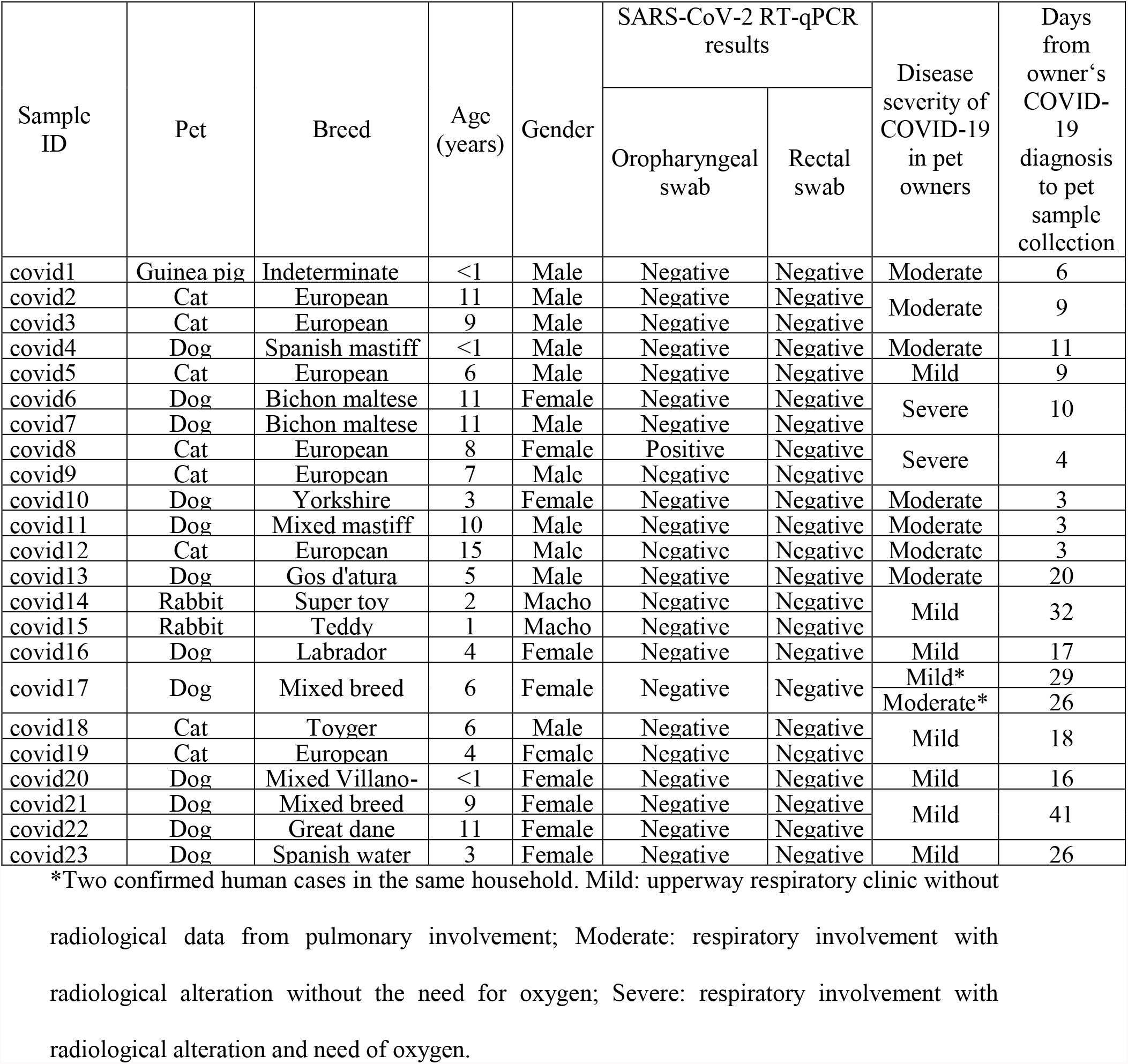
Characteristics of pets screened for SARS-CoV-2 living with owners diagnosed with COVID-19 during the quarantine (April-May 2020), in La Rioja (Spain).

Samples were aliquoted in a class II biological safety cabinet at a BSL-2 facility, and kept at −80°C. Carrier RNA (1μg) (Qiagen, Hilden, Germany) was added to each 200 μL-sample used for RNA extraction with RNeasy Mini Kit (Qiagen, Hilden, Germany). RNA extracts were eluted in 65 μL of RNAse-free water and stored at –80°C before use. DNA was digested using the RNase-Free DNase Set (Qiagen, Hilden, Germany). All samples were screened for SARS-CoV-2 using a specific one-step RT-qPCR assay targeting a fragment gene encoding the nucleocapsid (N) of the virus (2019-nCoV_N1 primers and probe set) [4] with the One Step PrimeScript™ RT-PCR Kit (Takara Bio Inc., Japan). A 20 μL reaction was set up containing 5μL of RNA extract, with 500 nM each primer and 125 nM probe. Thermal cycler was performed at 42 °C for 5 min for reverse transcription, followed by 95°C for 10 s and 45 cycles of 95 °C for 5s and 55° C for 30s. All specimens were also analysed using the commercial kit GPS™ CoVID-19 dtec-RT-qPCR Test (Genetic PCR Solutions™, Alicante, Spain). Positive cases were confirmed by RT-qPCR for the envelope (E) protein-encoding gene [5], as described above for the N1 gene, with these modifications: final concentrations of each primer and probe were 400 and 200 nM, respectively, and the annealing took place at 58°C for 30s. Synthetic plasmid controls with the complete SARS-CoV-2 N gene (Integrated DNA Technologies, Leuven, Belgium) and the E gene (Eurofins Genomics, Ebersberg, Germany) were used to generate standard curves based on ten-fold serial dilutions for quantification. The GPS™ CoVID-19 dtec-RT-qPCR Test kit also included a positive control. Positive and negative (extraction and amplification) controls were included in all the RT-qPCR assays. Pet samples and controls were tested in triplicate.

## Results and Discussion

From the 23 companion animals studied, the oropharyngeal swab sample from a female cat (ID covid8; Table) tested positive for SARS-CoV-2 by the three RT-qPCR assays performed. The specimen showed a viral load of 1.7×10^3^ RNA copies/μL for the N1 fragment gene, and 1.1×10^3^ RNA copies/μL for the E gene. Viral RNA was not detected in the rectal swab sample from this animal. It was an 8-year-old female domestic European cat that did not show clinical signs related to coronavirus disease, although it had chronic feline gingivostomatitis, feline idiopathic cystitis (treated with glucosamine and chondroitin sulphate), chronic kidney disease (treated with special feeding, ranitidine and benazepril hydrochloride) and feline asthmatic bronchitis (treated with fluticasone propionate). It lived in a two-cat household with a 7-year-old male European domestic cat that was negative for SARS-CoV-2 RT-qPCR assays from both swabs. Nevertheless, the efficient replication and transmission of the virus in cats has been experimentally demonstrated [6]. Follow up nasopharyngeal and rectal swab samples were subsequently taken on May 4, 2020 (E) protein-encoding gene(26 days later after the first sample collection) and both specimens from the two cats tested negative. The owner suffered from severe pneumonia and he was hospitalized for 8 days. Swab samples from the remaining mammalian pets screened for SARS-CoV-2 were negative.

Our study reports for the first time the detection of an asymptomatic cat with SARS-CoV-2 in Spain, probably associated with close contact with its owner who was diagnosed with active COVID-19 infection. According to our data, a high prevalence of SARS-CoV-2 RT-qPCR positive cats was observed (1/8; 12.5%). To date, the only prevalence data about animals published worldwide are those from Zhang et al. [7]. They found a 14.7% seroprevalence (but no molecular diagnosis) against SARS-CoV-2 in cats in China. To our knowledge, this is the first European case of SARS-CoV2 RNA detection in an asymptomatic cat, and the second reported in the world, after notification of one similar event in dogs in Hong Kong [8]. The rest of SARS-CoV-2 infections in felines worldwide (Belgium, USA, France and Spain) were symptomatic and mainly showed clinical signs of respiratory and/or digestive disease [9,10,11,12,13]. Probably, all these cases are also the result of contagions from their owners. Based on these results, it is possible that the number of affected cats living with COVID-19 owners is greater than that published to date, since these animals may be asymptomatic and not detected.

We consider that the limited number of animals included in our study can be a bias for the results. Nevertheless, the exceptional circumstances lived in Spain and in our region (La Rioja) during the sampling period (lockdown, confinement, quarantine, healthcare staff’s work overload, high mortality rates and health system on the brink of collapse) make these data relevant. Our finding suggests that cats may act as asymptomatic dispersers of the virus, although the transmission of SARS-CoV-2 from animals to humans seems unlikely [2]. All cases seem to be related with human transmission through symptomatic or asymptomatic COVID-19 infected people taking care of the animals. So far, all positive findings in pets were ‘isolated cases’ associated with close contact with humans positive for SARS-CoV-2, circumstance already described in the SARS epidemic in 2002–2003, without any epidemiological significance, considering them as dead-end hosts [14].

As a universal standard, hygiene measures should be exercised when living with a pet, especially if an infection is suffered. Currently, CDCs recommend restricting contact of people infected with COVID-19 with their companion animals [15]. At the same time, the OIE [16] highly recommends to keep positive animals isolated from other unexposed ones.

## Data Availability

The authors declare the availability of all data included in the manuscript

## Ethical statement

Institutional review board approval for this study was obtained from the Ethical Committee of Research with medicines from La Rioja (CEImLAR), (Ref. CEImLAR P.I. 419).

## Authors’ contributions

IRA: Ignacio Ruiz Arrondo; AP: Aránzazu Portillo; AMP: Ana María Palomar; SS: Sonia Santibáñez; PS: Paula Santibáñez; CC: Cristina Cervera; JAO: José A. Oteo.

IRA and JAO conceived and coordinated the study. IRA, AMP, SS, PS, CC and AP carried out the methodology. AP, IRA and AMP wrote the original draft. All authors reviewed, modified and approved the final version of the manuscript.

## Acknowledgements

We would like to acknowledge to the pets’ owners for their consent to include their pets in the study.

